# Ambulatory Blood Pressure and Number of Subclinical Target Organ Injury Markers in Youth: The SHIP AHOY Study

**DOI:** 10.1101/2024.03.15.24304137

**Authors:** Gilad Hamdani, Elaine M. Urbina, Stephen R. Daniels, Bonita E. Falkner, Michael A. Ferguson, Joseph T. Flynn, Coral D. Hanevold, Julie R. Ingelfinger, Philip R. Khoury, Marc B. Lande, Kevin E. Meyers, Joshua Samuels, Mark Mitsnefes

## Abstract

**Background:** Hypertension in adolescence is associated with subclinical target organ injury (TOI). We aimed to determine whether different blood pressure (BP) thresholds were associated with increasing number of TOI markers in healthy adolescents.

**Methods:** 244 participants (mean age 15.5±1.8 years, 60.1% male) were studied. Participants were divided based on both systolic clinic and ambulatory BP (ABP), into low- (<75^th^ percentile), mid- (75^th^-90^th^ percentile) and high-risk (>90^th^ percentile) groups. TOI assessments included left ventricular mass, systolic and diastolic function, and vascular stiffness. The number of TOI markers for each participant was calculated. A multivariable general linear model was constructed to evaluate the association of different participant characteristics with higher numbers of TOI markers.

**Results:** 47.5% of participants had at least one TOI marker: 31.2% had one, 11.9% two, 3.7% three, and 0.8% four. The number of TOI markers increased according to the BP risk groups: the percentage of participants with more than one TOI in the low-, mid-, and high groups based on clinic BP was 6.7%, 19.1%, and 21.8% (p=0.02), and based on ABP was 9.6%, 15.8%, and 32.2% (p<0.001). In a multivariable regression analysis, both clinic BP percentile and ambulatory SBP index were independently associated with the number of TOI markers. When both clinic and ABP were included in the model, only the ambulatory SBP index was significantly associated with the number of markers.

**Conclusion:** High SBP, especially when assessed by ABPM, was associated with an increasing number of subclinical cardiovascular injury markers in adolescents.

## Introduction

Cross-sectional studies have demonstrated that hypertension in childhood and adolescence is associated with subclinical markers of target organ injury (TOI).^1–4^ The Study of Hypertension in Pediatrics, Adult Hypertension Onset in Youth (SHIP AHOY) is a cross-sectional cohort study designed to determine BP levels and phenotypes that predict BP-related TOI in adolescents.^5^ Previous findings from this study have shown that BP levels below the current threshold used to define HTN were associated with individual measures of TOI (e.g., increased left ventricular mass abnormal diastolic function and left ventricular strain, and increased pulse wave velocity).^2,6,7^

A study in a pediatric chronic kidney disease (CKD) cohort from Europe found that systolic BP (SBP) level was independently associated with the number of intermediate TOIs found in these patients.^8^ However, whether the level of clinic BP and/or ABPM relates to severity of TOI as measured by the number of TOI abnormalities in a general pediatric population, is not known. In the current analysis, using data from the SHIP AHOY study we sought to determine whether increasing BP levels would be associated with increasing number of subclinical cardivascular TOI markers. We hypothesized that participants with higher BP would have a greater number of subclinical TOI markers.

## Methods

### Study Population

The design and methods of the SHIP AHOY study have been previously published.^5^ Briefly, in this cross-sectional study, youth aged 11 to 19 years were recruited at six clinical sites (Cincinnati, Houston, Philadelphia, Rochester, Seattle, and Boston) from 2015 to 2018. Youth with symptomatic severe hypertension, on antihypertensive or lipid-lowering medication in the past 6 months, with diabetes mellitus (type 1 or 2), kidney diseases, or other chronic medical conditions were excluded. All study participants and their parents provided written informed consent and assent according to local investigational review board requirements. In the current analysis, only data from participants who completed both an ABPM and an assessment of all four measures of TOI listed below were included.

Patient demographic data were self-reported by each participant. Measures of height and weight were obtained with calibrated scales and stadiometers. Body mass index (BMI) was calculated as weight/height^2^. Laboratory analyses were performed on fasting serum samples.

### Clinic BP

Seated BP measurements were obtained in the right arm by auscultation using a calibrated aneroid sphygmomanometer (Mabis MedicKit-5; Mabis Healthcare, Waukegan, IL) as previously described.^5^ Four BP measurements at 30-second intervals were obtained on each of 2 separate study visits, with the average of the second, third, fourth and sixth, seventh, and eighth measurements recorded as the clinic BP value. SBP percentile was calculated based on the 2017 American Academy of Pediatrics guidelines.^9^ For this analysis, participants were divided into 3 at-risk subgroups based on their clinic SBP percentile: low-risk <75^th^ percentile; mid-risk 75^th^-90^th^ percentile, and high risk >90^th^ percentile.

### ABP Measurements and Classification

Ambulatory BP was measured according to published pediatric guidelines,^10^ as previously described using the OnTrak 90227 device (SpaceLabs, Snoqualmie, WA), an oscillometric BP monitor that uses the same algorithm as the device that was used to generate the most commonly used pediatric normative ABPM data set.^15^ ABPM was performed per American Heart Association (AHA) guidelines with specific attention to proper cuff sizing according to arm circumference.

Only ABPM records consistent with current AHA recommendations for ABPM and pediatric ABPM guidelines (at least 65-75% of successful readings with > 40 readings available in 24-hour period) were included in the analysis.^10,11^Awake mean SBP percentile was calculated according to pediatric normative data.^12^ Similar to how we categorized clinic BP, participants were divided into 3 at-risk subgroups based on mean awake SBP percentile: low-risk ABP, <75^th^ percentile; mid-risk ABP, 75^th^ – 90^th^ percentile, and high-risk ABP, >90^th^ percentile.

### Target Organ Injury (TOI)

Methodology for evaluation of TOI in SHIP AHOY has been previously described.^5^ Standard measurements of left ventricular mass, systolic function, and diastolic function were obtained from echocardiograms. Images were read by an experienced sonographer using the Cardiology Analysis System (Digisonics, Houston, TX):

1. Left ventricular mass was calculated using the Devereux equation from 2-dimensional M-Mode images of the left ventricle at end diastole.^13^ The left ventricular mass index (LVMI) was defined as left ventricular mass/height^2.7^ as described by DeSimone et al^14^ to account for body size without overcompensating for obesity. Consistent with previous SHIP AHOY analyses,^6^, LVH was defined as LVMI >38.6 g/m^2.7^, the 95^th^ percentile of LVMI for adolescents.^15^
2. Systolic function was evaluated by tracing the endocardium from the 4-chamber view at peak systole and end diastole (TOMTEC Corporation, Chicago, IL) to quantify global longitudinal strain (GLS), strain rate, time to peak longitudinal strain, time to peak longitudinal strain rate, and left ventricular ejection fraction.^16^ GLS measurements were obtained at native echo frame rates (40–60 frames per second). Abnormal systolic function was defined as GLS >-18.^17^
3. Diastolic function was assessed using pulse wave Doppler of the mitral inflow velocity in the apical 4-chamber view to determine E/A ratio. Tissue Doppler imaging of the mitral annular inflow was recorded at the lateral and septal annulus with the e’/a’ and E/e’ ratios from both regions averaged. Abnormal diastolic function was defined as E/e’ >8.^18^

Pulse wave velocity (PWV), a marker of vascular stiffness, was noninvasively measured using the SphygmoCor CPV System (AtCor Medical, Sydney, Australia).^19,20^ Carotid-femoral distance was calculated as the sum of the distance from the suprasternal notch to the femoral artery pulse (using a caliper) minus the distance from the carotid pulse to the suprasternal notch (measured with a tape measure) according to accepted guidelines.^19^ These measurements were made twice, averaged, and entered into the device. The pulse transit time is determined by the device as the difference in time between the peak of the R-wave (from the applied ECG leads) to the foot of the femoral pressure wave (obtained with a tonometer) minus the R-wave to the foot of the carotid pressure wave time. PWV is calculated as distance divided by difference in time. PWV is highly reproducible with coefficient of variation on replicate measures of 7%.^21^ Based on the 90^th^ percentile of normotensive participants in this cohort and lean adolescents in a previous study^22^, elevated arterial stiffness was defined as PWV>5.8 cm/sec.

For each participant, we summed the number of TOI assessments that fulfilled the above criteria.

### Statistical Analyses

For descriptive statistics, categorical variables were reported as percentages, and continuous variables as mean ± SD or median (IQR). Number of abnormal TOI measures, and percentage of patients with >1 TOI was compared between groups by χ2 analyses, while differences in continuous variables were evaluated by ANOVA. General linear models with the number of TOI markers as the dependent variable and different SBP measures as the primary independent variable were performed. For the multivariate analysis, all parameters collected were initially included in the model. Backward elimination was performed to determine variables included in the formal model, with an inclusion criterion of *p*<0.05. All analyses were performed using SAS^®^ 9.4 for Windows (SAS Institute Inc., Cary, NC).

## Results

### Study Population

Study participant characteristics are detailed in **Table 1**. Of the 397 participants of the SHIP AHOY study, 244 completed a full assessment of all TOI markers and ABPM. Mean age was 15.5 years, 60.1% were male and 66.8% were white. Mean height was 169.2±10.9cm, and mean weight was 80.1±25.2 kg. The study population was relatively overweight with a mean BMI percentile of 80.7±25.4 and tended towards insulin resistance with a mean HOMA_IR of 4.6±4.3.

**Table 1:** Study Participant Characteristics.

| **Variable (N=244)** | **Mean ± SD or Frequency** |
| --- | --- |
| Age (years)* | 15.5±1.8 |
| Race, n (%) |  |
| White | 163 (66.8) |
| Black | 58 (23.8) |
| Asian | 11 (4.5) |
| Other | 12 (4.9) |
| Sex, males, n (%) | 146 (60.1) |
| Height (cm) | 169.2±10.2 |
| Weight (kg) | 80.1±25.2 |
| BMI (kg/m<sup>2</sup>) | 27.7±7.4 |
| BMI percentile (%) | 80.7±25.4 |
| Total cholesterol (mg/dl) | 153.6±32.9 |
| LDL-C (mg/dl) | 89.8±27.6 |
| HDL-C (mg/dl) | 45.3±12.3 |
| Triglycerides (mg/dl) | 97.4±55.0 |
| Glucose (mg/dl) | 89.9±9.2 |
| Insulin (microIU/ml) | 20.3±17.5 |
| Creatinine (mg/dl) | 0.7±0.2 |
| Uric acid (mg/dl) | 5.7±1.6 |
| hsCRP (mg/dl) | 1.6±1.9 |
| Urine sodium/potassium ratio | 3.9±2.7 |
| HOMA_IR | 4.6±4.3 |
| TG/HDL ratio | 2.5±2.3 |
| SBP (mmHg) | 122.6±12.8 |
| DBP (mmHg) | 71.4±10.6 |
| SBP percentile (%) | 75.5±27 |
| DBP percentile (%) | 64.4±28.7 |
| HR (beats/min) | 71.1±12.5 |
| ABP wake SBP (mmHg) | 124.4±11.3 |
| ABP sleep SBP (mmHg) | 108.6±10.5 |
| ABP wake DBP | 72.0±7.8 |
| ABP sleep DBP | 58.2±7.4 |
| LVM Index (g/m<sup>2.7</sup>) | 32.2±7.0 |
| E/A ratio | 2.2±0.6 |
| E/e' ratio | 6.2±1.5 |
| e'/a' ratio | 2.4±0.7 |
| LV longitudinal strain (%) | -20.4±3.5 |
| Shortening Fraction (%) | 37.4±4.6 |
| Pulse wave velocity (m/sec) | 5.1±0.9 |

### Blood Pressure Status

According to clinic BP, 75 participants (30.7%) were included in the low-risk BP group, while 68 (27.9%) and 101 (41.4%) were included in the mid- and high-risk groups, respectively. Participant characteristics based on clinic SBP category are detailed in **Supplementary Table 1**. Participants in the high-risk BP group had higher glucose, uric acid, and hsCRP serum levels compared to the low-risk group, and higher insulin and HOMA-IR levels compared to the mid-risk group.

Based on ABPM results, 135 participants (58.2%) were included in the low-risk ABP group, while 38 (16.4%) and 59 (25.4%) were included in the mid-and high-risk groups, respectively. Participant characteristics based on awake ambulatory SBP category are detailed in **Supplementary Table 2**. Participants in the high-risk ABP group were heavier, had higher waist circumference, waist to height ratio, BMI, and BMI percentile than those in the other groups. Participants in this group also had lower HDL-cholesterol and higher hsCRP levels.

**Table 2:**
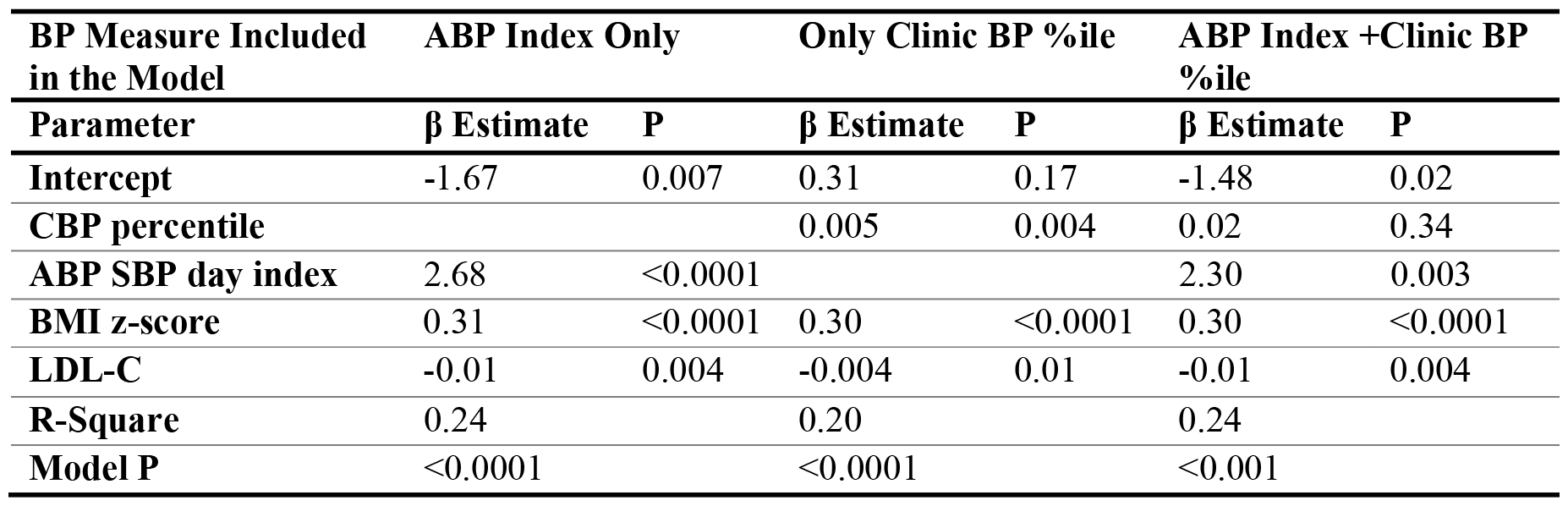
Independent Correlates of the Number of TOI Abnormalities.

| **BP Measure Included**<br>**in the Model** | **ABP Index Only** |  | **Only Clinic BP %ile** |  | **ABP Index +Clinic BP**<br>**%ile** |  |
| --- | --- | --- | --- | --- | --- | --- |
| **Parameter** | **$\beta$ Estimate** | **P** | **$\beta$ Estimate** | **P** | **$\beta$ Estimate** | **P** |
| **Intercept** | -1.67 | 0.007 | 0.31 | 0.17 | -1.48 | 0.02 |
| **CBP percentile** |  |  | 0.005 | 0.004 | 0.02 | 0.34 |
| **ABP SBP day index** | 2.68 | <0.0001 |  |  | 2.30 | 0.003 |
| **BMI z-score** | 0.31 | <0.0001 | 0.30 | <0.0001 | 0.30 | <0.0001 |
| **LDL-C** | -0.01 | 0.004 | -0.004 | 0.01 | -0.01 | 0.004 |
| **R-Square** | 0.24 |  | 0.20 |  | 0.24 |  |
| **Model P** | <0.0001 |  | <0.0001 |  | <0.001 |  |

### Subclinical Target Organ Injury

One hundred and sixteen participants (47.5%) had at least one abnormal TOI marker: 76 (31.2%) with one marker of injury, 29 (11.9%) with two, 9 (3.7%) with three, and 2 (0.8%) with 4. The most common form of TOI marker was abnormal global longitudinal strain (23.7%), followed by increased PWV (17.7%), LVH (17.2%), and abnormal diastolic function (9.9%). Measures of TOI markers based on BP status are presented in **Supplementary Tables 1 and 2**. In general, measures LVMI, E/e’, and PWV were higher in the high-risk BP group, according to both clinic and ambulatory BP.

The percentage of participants with a specific sum (0,1,2,3, or 4) of subclinical TOI markers in each clinic and ambulatory BP risk group are presented in **Figures 1 and 2**, respectively. In both cases there was a stepwise increase in the number of TOIs between categories. For clinic BP, the percentage of participants >1 TOI marker in the low-, mid-, and high-risk groups was 6.7%, 19.1%, and 21.8% BP (p=0.02), and for ABP it was 9.6% 15.8%, and 32.2%, respectively (p<0.001).

**Figure 1:**
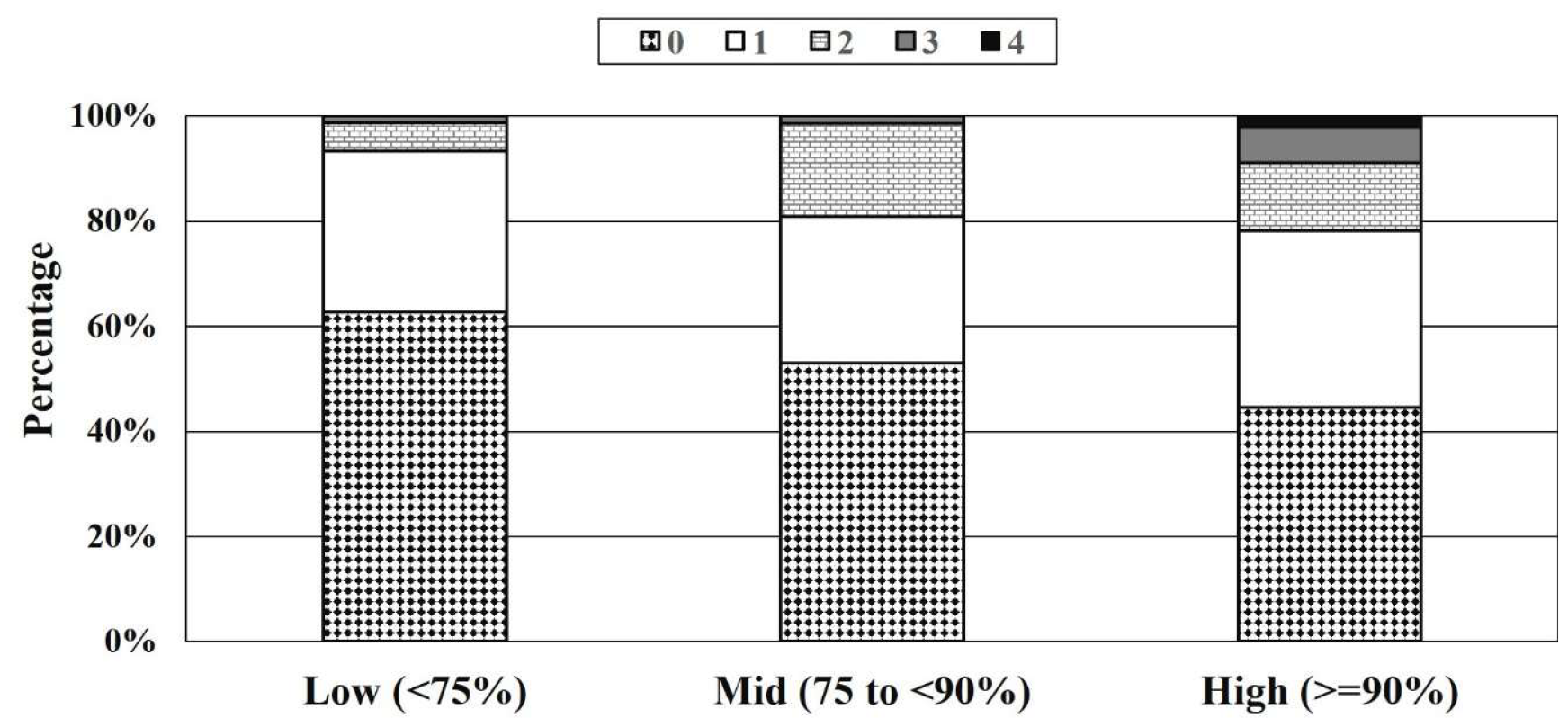
Percentage of Participants with TOI According to Clinic SBP Categories.

**Figure 2:**
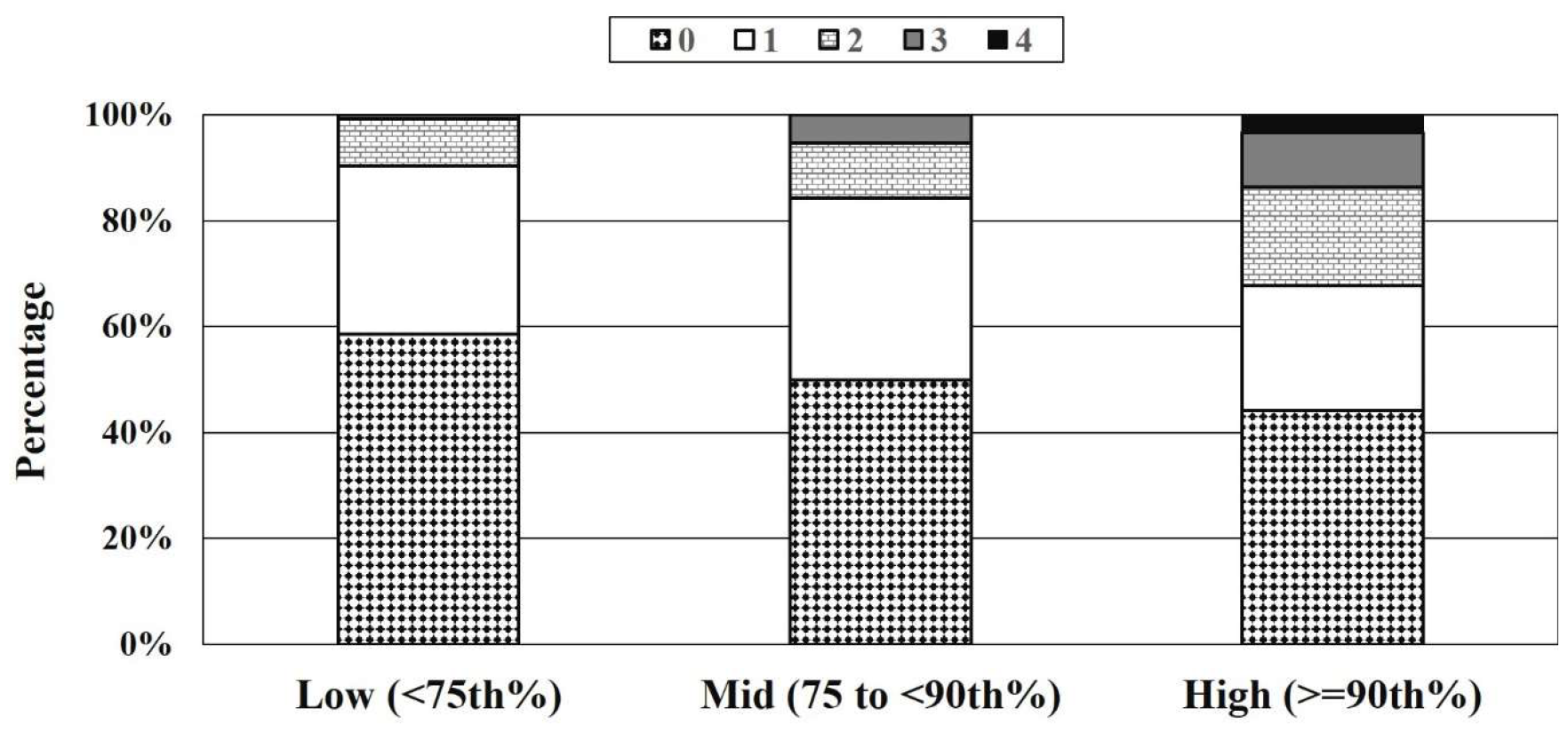
Percentage of Participants with TOI According to Ambulatory SBP Index Categories.

In a multivariable regression analysis **(Table 2)**, both clinic SBP and awake ambulatory SBP were independently associated with the number of TOI markers. When included in the same model, only ambulatory SBP index was significantly associated with the number of TOI markers. A regression analysis using absolute clinic BP values and mean wake ambulatory SBP values yielded similar results (**Supplementary Table 3**).

## Discussion

In this cohort of otherwise healthy adolescents, one-third of participants had one or more marker of TOI, and the number of TOI markers increased with increasing clinic and ambulatory SBP. We also determined that this association was stronger with ABP rather than clinic BP. This is consistent with previous studies in both adults and children showing that ABP is a superior predictor of individual clinical and intermediate cardiovascular outcomes.^23–27^

A recent study from the International Childhood Cardiovascular Cohort (i3C) Consortium found that a combined risk score comprised of several traditional cardiovascular risk factors better predicted cardiovascular events adulthood than each risk factor by itself.^28^ In that study, 20,656 participants who had cardiovascular risk factors measured in childhood had their vital status ascertained in adulthood. Each of the risk factors that were available for each child significantly predicted fatal and non-fatal events: hazard ratio (HR) for BMI 1.45, total cholesterol 1.31, triglycerides 1.45, smoking 1.70, SBP 1.33. However, the combined risk score had the highest HR that persisted after correction for demographics, year of ascertainment and cohort (2.75, 95% CI 2.48-3.06). This suggests a combined risk score is more powerful in predicting later outcomes. Our study adds to the literature by demonstrating that in addition to multiple traditional risk factors, adolescents with high BP may already have multiple markers of TOI, long before the age at which BP-related cardiovascular events have been described to occur. This is relevant, as these measures of TOI in youth are associated with future adult cardiovascular diseases.^29,30^ Identifying adolescents with TOI earlier in life might allow for more effective preventive measures.

Our study has several limitations. First, the participants in our study were relatively overweight and thus might not represent the general adolescent population, especially since being overweight by itself is associated with adverse cardiovascular outcomes. On the other hand, our cohort is likely representative of the typical adolescent population referred for hypertension evaluation.^31^ To address this limitation, all our analyses were adjusted for BMI. The cross-sectional analysis precludes determination of causality between the different BP measures and TOI markers. As our cohort included generally healthy adolescents, associations with more robust cut-offs of TOI, such as higher levels of LVMI and E/e’, were impractical. We therefore used lower cut-points that have previously been used in similar populations. Strengths of the present study include the careful measurement of both clinic and ambulatory BP using gold-standard procedures across multiple sites, as well as the use of state-of-the-art assessments of target organ consequences of exposure to high BP.

### Perspective

Our findings demonstrate that high SBP, even at levels below HTN thresholds, was associated with an increasing number of subclinical cardiovascular injuries in adolescence, especially if ABPM is used to assess BP status. These findings highlight the importance of BP monitoring in adolescence, and the application of ABPM if elevated BP levels are observed. Further research is needed to establish this association as well as the potential significance of the number of subclinical abnormalities on cardiovascular events in later life.

### Novelty and Relevance

What Is New?

- High systolic BP is not only associated with specific markers of subclinical cardiovascular target organ injury in adolescence, but with an increasing number of several such markers.
- The association of ambulatory PB with increasing number of target organ injury markers is stronger than that of clinic blood pressure.

What Is Relevant?

- Our study demonstrates the strength of the association between high SBP, even at levels below hypertension thresholds, and several subclinical markers of cardiovascular target organ injury, even at an early age.

Clinical/Pathophysiological Implications?

- BP monitoring in adolescence is important.
- Application of ABPM is needed if elevated BP levels are observed.
- Further research is needed to establish the potential significance of the number of subclinical abnormalities on cardiovascular events in later life.

## Data Availability

This choort's data is not available for public access.

## Sources of Funding

Supported by the American Heart Association funding (Grant #15SFRN23680000) to Cincinnati Children’s Hospital Medical Center and University of Cincinnati College of Medicine; and by NIH National Center for Advancing Translational Sciences grants to the University of Cincinnati (#UL1 TR001425), University of Washington (#UL1 TR002319), and University of Rochester *(#UL1 TR002001)*.

### List of Non-standard Abbreviations

*ABP*: Ambulatory Blood Pressure
*ABPM*: Ambulatory Blood Pressure Monitoring
*AHA*: American Heart Association
*CKD*: Chronic Kidney Disease
*GLS*: Global Longitudinal Strain
*LVH*: Left Ventricular Hypertrophy
*LVMI*: Left Ventricular Mass Index
*PWV*: Pulse Wave Velocity
*SBP*: Systolic Blood Pressure
*TOI*: Target Organ Injury

## Supplemental Material

Tables S1-3

